# Personalised medicine for Crohn’s disease is a cost-effective strategy

**DOI:** 10.1101/2022.12.01.22281309

**Authors:** Vanessa Buchanan, Susan Griffin, Warda Tahir, Karen Hills, Miles Parkes, Kenneth GC Smith, Paul A Lyons, James C Lee, Eoin F McKinney

## Abstract

**Objective:** To evaluate the cost-effectiveness of a personalised medicine strategy for Crohn’s disease in the UK, using early targeted top-down therapy compared to standard of care.

**Materials & Methods:** A decision tree leading into a Markov state-transition model was constructed, allowing comparison of two treatment approaches: 1) standard of care therapy following established UK clinical guidelines (‘step-up’ treatment) and 2) a personalised medicine strategy in which patients identified as high-risk of subsequent relapse using a prognostic biomarker receive ‘top-down’ anti-TNF treatment at diagnosis. The model facilitated comparison of both costs and Quality Adjusted Life Years (QALYs) in a hypothetical cohort of newly diagnosed Crohn’s disease patients with sensitivity analyses undertaken to model the impact of key assumptions.

**Results:** Early personalised treatment with anti-TNF based combination therapy resulted in an incremental cost-effectiveness ratio (ICER) of £2,176 per quality-adjusted life year (QALY), with £717 incremental costs and 0.330 incremental QALYs, substantially below the NICE cost-effectiveness threshold of between £20,000 and £30,000 per QALY. Additional costs relating to earlier biologic use were offset by incremental QALYS and reductions in costs driven by fewer disease flares and hospitalisations. Sensitivity analysis across a wide range of parameter assumptions did not impact on the model’s conclusion.

**Conclusion:** A personalised medicine strategy using anti-TNF therapy at diagnosis in Crohn’s disease to patients at high risk of subsequent relapse is highly likely to be a cost-effective use of resources in the UK National Health Service.

**KEY SUMMARY:** *Established Knowledge:*

- Currently there are no validated prognostic test that can stratify IBD patients based on long term outcomes at the point of diagnosis used routinely in the UK
- It therefore remains unclear which patients with Crohn’s disease should be treated with early anti-TNF based therapy as part of a ‘top-down’ regimen.
- As a consequence, the majority of IBD patients in the UK are currently treated with an accelerated step-up approach

*Significant new findings:*

- We show here that the use of biomarkers at diagnosis to guide personalised use of such treatment is a cost-effective approach for treatment of Crohn’s disease.
- Use of a prognostic test to deliver personalised medicine for Crohn’s disease results in positive QALY of 0.330
- The approach is cost effective with an incremental cost of £717 and an ICER of 2,176
- The model’s conclusions were unaffected by a wide range of sensitivity analyses

## INTRODUCTION

Clinical outcomes in inflammatory disease such as Crohn’s Disease (CD) are heterogeneous, creating uncertainty in treatment decisions as two patients with similar clinical presentations may each experience very different disease courses. Without being able to predict clinical outcome, a personalised medicine strategy – in which treatment is matched to patients destined for an aggressive disease course – is not possible. Such clinical biomarkers have recently become available, but in order to achieve widespread acceptance it must be demonstrated that they can not only reliably predict disease course^1^ but also that the information provided can improve both efficacy^2^ and efficiency of treatment regimens. The latter requires modelling their impact on the cost-effectiveness of current therapy guidelines.

National and International guidelines, including from the European Crohn’s and Colitis Organisation (ECCO) and the British Society of Gastroenterology (BSG), recommend a treatment strategy for CD that is reactive, with stepwise escalations in therapy being used in response to recurrent flares or persistently active disease^2^. The UK National Institute for Health and Care Excellence (NICE) guidelines state that patients with CD should initially be treated with corticosteroids to induce remission, and that if the symptoms worsen or the disease is resistant to corticosteroids then therapy should be incrementally escalated, with the addition of immunosuppressive medications and biologic therapy (‘step-up’ therapy). Biologic therapy, including anti-TNFα, is only recommended for people with severe active disease which has not responded to conventional therapy, or where conventional therapy is not tolerated for maintaining remission^3^.

However, early use of anti-TNF therapy as part of a ‘top-down’ treatment strategy has been repeatedly shown to improve clinical outcomes in CD compared to standard therapy^4–7^. This approach also maximises the potential to achieve disease remission since delayed introduction of anti-TNF has been consistently shown to reduce response rates. Early biologic therapy can also deliver savings from reduced disease-related adverse events such as hospitalisation, flares and surgery^8, 9^. However, increasing early use of more potent biologic treatments inevitably impacts on the cost-effectiveness of any treatment: it must be effective enough to outweigh any associated risks and costs, in order to have a net improvement on the treatment strategy. Untargeted use of top-down treatment strategies have not gained widespread adoption, due to the inevitable exposure of low-risk patients to the toxicity of avoidable immunosuppression and the increased healthcare costs associated^10^. The ability to reliably predict an individual’s clinical outcome at the point of diagnosis^1, 10^ provides the maximum “window of opportunity” to harness the benefit of top-down therapy while minimising the associated risk and costs. In effect, personalising therapy can improve cost-effectiveness of therapy by only exposing patients who stand to gain most from them.

Reduction in healthcare associated costs is a major factor in the adoption of new healthcare technology and consequently in the translation of personalised medicine from the bench to the bedside in Crohn’s disease: it is not enough for new treatments to demonstrate efficacy as, for adoption into clinical use, impact on cost-effectiveness is also required^11, 12^. Here, we formally test the cost-effectiveness of a personalised medicine strategy in Crohn’s disease in which use of top-down treatment (comprising combination or monotherapy with anti-TNF therapy) at diagnosis is guided by a prognostic marker. We compare healthcare costs (from drugs, hospitalisations and complications) with benefits (incremental QALYs) resulting from both the personalised strategy and the current standard of care (as per UK NICE guidelines). We go further to investigate the impact of key modelling parameters on the cost-effectiveness of a personalised approach, testing the sensitivity of our conclusions to assumptions of treatment efficacy, biomarker performance, drug costs and model time horizon.

## MATERIALS & METHODS

A decision tree leading into a Markov state transition model was designed to evaluate the cost-effectiveness of biomarker-guided top-down therapy compared with standard of care (SoC) step-up therapy in newly diagnosed CD patients. The model captured the frequency of flares (defined as episodes of active CD requiring escalated immunomodulator therapy), hospitalisations and surgery and calculated both the costs and QALYs associated with the two treatment approaches in a hypothetical cohort of newly diagnosed CD patients. Model assumptions, patient characteristics and biomarker performance were informed by prior evidence as cited and guided by independent expert review.

### Population, Interventions and Comparators

The model population was treatment-naïve patients with newly diagnosed CD. Patients eligible for immediate top-down therapy (complex perianal disease, significant fistulising disease or multiple risk factors) were excluded from the personalised strategy as there is a clear indication for early biologic treatment. The NICE technology appraisal (TA) of ustekinumab for the treatment of CD^13^ was used to generate representative baseline patient characteristics^14^ as presented in Table 1.

**Table 1:** Baseline characteristics of patients entered in the model.

| Characteristic | Value |
| --- | --- |
| Starting age (years) | 40 |
| Proportion with high-risk CD | 55% |
| Mean patient weight (kgs) | 72 |
| % patients <55kg | 21% |
| % patients 55kg<x<85g | 59% |
| % patients >85kg | 20% |

Patients were allocated to one of two treatment approaches (Figure 1):

**Figure 1.**
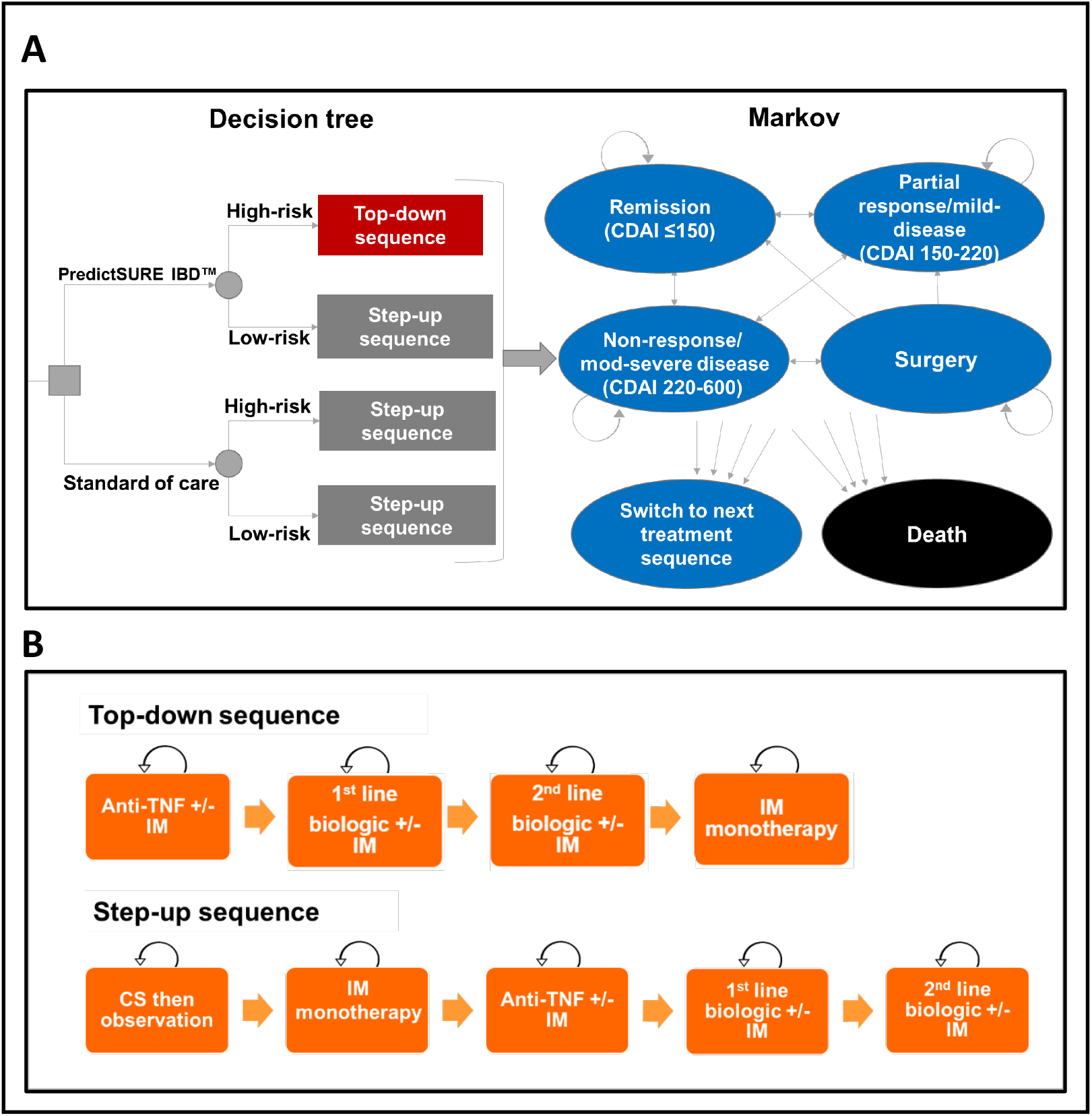
Model structure. **A**, Model structure diagram demonstrating the decision tree and Markov components; No patients were assumed to have a CDAI score greater than 450 (very severe disease) until the end of sequence, whereupon 50% of patients were assumed to have very severe disease **B,** Step-up and Top-down treatment sequences CDAI, Crohn’s disease activity index; IM, immunomodulator; CS, corticosteroid

#### Personalised therapy

The intervention in the personalised therapy group was targeted anti-TNF therapy guided by biomarker assay results^1^, whereby patients identified as high risk receive anti-TNFα therapy combined with an immunomodulator, followed by other biologic classes upon relapse (‘top-down’ treatment, TD):

1) Induction with anti-TNF therapy (allocated to patients as 80% adalimumab, 20% infliximab) and immunomodulator (allocated to patients as 80% azathioprine, 10% 6-mercaptopurine and 10% methotrexate). Proportions of treatments were informed by clinical opinion.
2) Second line biologic, allocated as 50% vedolizumab and 50% ustekinumab
3) Third line biologic, allocated as 50% vedolizumab and 50% ustekinumab (converse of selection used in step 2)
4) Immunomodulator alone (allocated as per step 1)

Time to event data for the personalised therapy subgroups (high and low risk) was based on the validation study of the PredictSure-IBD prognostic assay^1^ as indicated in Figure 2A.

**Figure 2:**
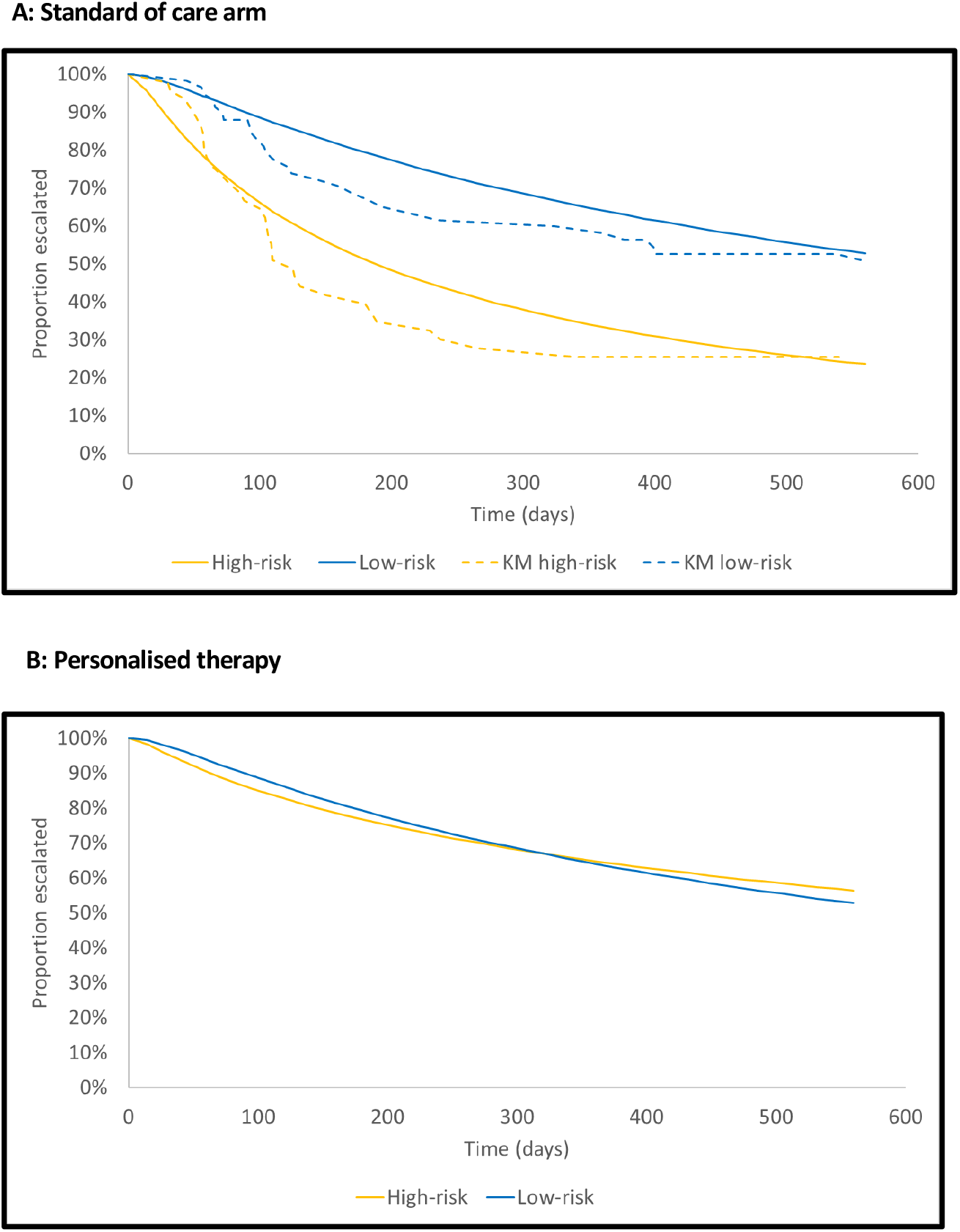
*Kaplan-Meier* survival curves showing time to first treatment escalation for each model arm. **A,** Comparison of the lognormal parametric survival curves (solid lines) fitted to the Kaplan-Meier time to escalation patient data (dotted lines), stratified by risk group. **B,** Treatment effect of using a TD therapy approach on high-risk patients, modelled by applying a hazard-ratio of 0.53 to the high-risk fitted time to escalation curve. KM, Kaplan-Meier survivor function.

#### Standard care

The standard care group received treatment following established UK clinical guidelines^15^, consisting of sequences of immunomodulator followed by biologic upon relapse (‘step-up’ treatment, SU):

1) Treatment with corticosteroid (prednisolone), followed by observation until relapse
2) Immunomodulator (as per TD)
3) anti-TNF therapy and immunomodulator (as per TD)
4) A second-line biologic allocated as 50% vedolizumab and 50% ustekinumab
5) A third-line biologic allocated as 50% vedolizumab and 50% ustekinumab (converse of selection used in step 4)

The biomarker assay used was modelled on the performance of PredictSure-IBD with assay performance evaluated in an independent cohort as described^1^. The PredictSure-IBD assay was used as an exemplar for the impact of personalised medicine, although broader relevance to other putative predictive models was facilitated through sensitivity modelling of the model’s result to key performance characteristics including treatment effect size on subgroups identified.

### Model structure

A decision tree leading into a Markov state-transition model was constructed in Microsoft Excel to compare personalised and standard therapy approaches. The decision tree component allocates patients to a clinical prognostic group (high or low risk) and to a treatment strategy (personalised or standard therapy). Patients in both the personalised and standard treatment groups were similarly allocated a prognostic group, the only difference being that this information is ‘known’ in advance in the personalised therapy group with treatment assigned accordingly with high risk individuals receiving top-down therapy and low-risk individuals receiving standard therapy. The ‘standard’ group comprises identical % of individuals with high and low risk outcomes to the personalised group, the only difference being that both groups receive the same initial therapy (following current guidelines) as the outcome is not known in advance (Figure 1A).

Patients in each prognostic group and treatment strategy then enter the Markov phase of the model. A Markov model is comprised of a set of mutually exclusive and exhaustive health states that represent disease progression, in this case CD severity score as measured on the CD activity index (CDAI). Health states were defined as Remission (CDAI≤150), Partial response/mild disease (CDAI 150-220), Non-response/moderate-severe disease (CDAI 220-600), Surgery or Death. The health states capture the costs and health-related QoL (utility) scores associated with each level of severity and any adverse events that occur as a result of either the disease or therapy^16^. A Markov-based analysis was selected as this model structure is most suited for chronic diseases such as CD, with features such as the ability to incorporate temporal detail such as recurrent events^17^ which other model structures do not allow for.

As step-up and top-down therapy both comprise a series of discrete treatments, the Markov component of the model was constructed to model treatment sequences, with each treatment divided into induction and maintenance phases. While on treatment, in order to reflect the relapsing-remitting nature of CD, patients can cycle through the three CDAI health states, can have a disease flare and can undergo surgery. The Markov cycle length was 14 days with a time horizon of 20 years. Parameters were chosen to reflect both the relapsing-remitting and chronic nature of CD balanced against inherent uncertainty of extrapolating evidence-based assumptions across increasing time horizons and are in line with previous cost-effectiveness models in chronic relapsing disease^3, 13, 18, 19^. Additional sensitivity modelling was undertaken to test their impact on model outcome as detailed below.

Patients escalate from one treatment to the next through the defined schedule (personalised or standard therapy) when they experience a disease flare requiring treatment escalation. Patients on TD have a reduced risk of experiencing adverse events, including surgeries and hospitalisations, compared to SU with an effect size estimated from prior studies and with sensitivity modelling as outlined below. Once treated, patients can experience mucosal healing (absence of ulcers) after which the probability of achieving clinical remission (CDAI<150) on treatment is increased. More patients in the model are assumed to achieve mucosal healing and hence remission on TD than on SU^8, 20^.

Once patients fail the last treatment in the sequence, they are allocated equally to the moderate and severe health states. Although in practice treatment-refractory patients may receive further investigational medicines, this was not explicitly modelled as clinical practice varies widely for such refractory cases. The model also incorporates an age-related decline in utility^21^.

### Assumptions

The key assumptions underlying the model are provided in table 2. Model assumptions were validated against key prior studies as cited and validated by independent, expert review.

**Table 2:** Key assumptions of Markov model.

| Assumption | Explanation |
| --- | --- |
| Treatment effect on time to escalations/flare (relapse) is 0.4, greater than that reported in D'Haens et al<sup>6</sup>, which was 0.53 | In the D'Haens trial, the median time to relapse was 174.5 vs 329 days for SU and TD respectively. Using these figures, we derived a hazard ratio on time to relapse of 0.53. However, it was assumed that this was an underestimate of the efficacy of top-down treatment as<br>the top-down group received infliximab as induction treatment only, and patients received maintenance azathioprine rather than continuing TNFα inhibitor maintenance therapy as per current guidelines<sup>29</sup>.<br>Therefore, we adjusted the treatment effect on time to relapse to reflect the greater efficacy that is known to result from combined Infliximab and azathioprine treatment. |
| Treatment effect of top-down on risk of flares, treatment escalations and surgery assumed to last 10 years | Effects on key clinical outcomes were observed up to 10 years after randomisation to the top-down vs. step-up treatment arms in D'Haens trial<sup>6</sup>, as reported in Hoekman et al<sup>23</sup> |
| After 12 months of maintenance biologic, 85% of step-up patients remain on regular maintenance within that step. | While NICE recommends ongoing use of biologic after 12 months based on clinical need, only 15-20% of patients in clinical practice discontinue biologics after 12 months, hence we assumed that approximately 85% would remain on therapy. |
| After 12 months of maintenance biologic, 20% of top-down patients remain on regular maintenance within that step | There is a paucity of evidence to support what proportion of patients given top-down biologic is likely to require treatment after 12 months, but due to better response this is likely to be significantly lower than in immunomodulator-refractory disease. Based on clinical expert opinion, it was assumed that only 20% of patients were likely to require treatment after 12 months. |
| The effect of TD on mucosal healing is assumed to be sustained over all treatments in the sequence | 76% and 40% of top-down vs. step-up patients respectively achieved mucosal healing after 6 months in the D'Haens trial<sup>6</sup>. The effect of mucosal healing on the proportion in remission (odds ratio of 4.3<sup>20</sup>) was found to be sustained for up to 4 years. Although long-term remission was not observed in the long-term follow-up of the D'Haens study<sup>23</sup>, this may reflect the use of biologic during induction only and lack of 2<sup>nd</sup> and 3<sup>rd</sup> line alternatives. The assumption was therefore made of longer-term impact on remission |
| The risk of flares not requiring treatment escalation was assumed to be constant over time | Due to small event rates, an exponential distribution was fitted to time to escalation-free flare, with the implication that the risk of flare is constant over the model lifetime |
| Risk of treatment escalation and/or surgery following the first event was assumed to be constant over time | Due to the Markovian memory limitation and diminishing patient numbers over the follow-up time, an exponential distribution was fitted to time to treatment escalation and time to surgery following the first event, with the implication that risk is constant over the remaining model lifetime |
| Distribution across CDAI health states conditional on treatment was assumed to remain constant over time | Due to the Markovian memory limitation, an exponential distribution had to be fitted to this parameter, with the implication that this parameter is constant over time |

### Model parameters

#### Clinical

The proportion of patients allocated to high/low risk subgroups and their respective clinical outcomes were obtained from pooled individual patient data from a prospective cohort of patients with active IBD, tested prior to initiation of therapy^1, 10, 22^.

The probabilities of treatment escalation, disease flare and surgery for each group were estimated by fitting parametric survival curves to the Kaplan-Meier time to event data of the cohort using STATA 14 (StataCorp) and extracting time-dependent transition probabilities. Choice of parametric function was determined by goodness of fit statistics and visual fit. The treatment effect (hazard ratio) of TD on time to treatment escalation and flares was based on median values reported in the “Step-Up Top-Down” trial^6^ with sensitivity modelling as described below. Note that treatment effect size is a hazard ratio of disease flare in the high-risk group with/without early anti-TNF therapy and consequently a smaller value reflects a larger treatment effect. The treatment effect of TD on surgery was calculated from the number of surgeries reported in the extended follow-up of the “Step-Up Top-Down” study^23^. The treatment effect of TD vs. SU on treatment escalation was assumed to wane over time until no treatment effect remained by 10 years (the duration of follow-up in the study on which the assumption is founded^23^). CDAI scores to inform allocation to disease severity health state were derived from prior studies and independent expert review^13, 19^.

Mean values for all clinical input parameters are provided in supplementary table S1, along with their respective distributions and sources.

### Quality-adjusted life years

The economic evaluation took the form of a cost-utility analysis with outcomes measured in quality-adjusted life years (QALYs). QALYs corresponding to the health states and decrements in QALYs for adverse events were extracted from multiple published studies, as detailed in Table S1. Outcomes were discounted at an annual rate of 3.5% after the first year^24^.

### Costs

The economic evaluation was conducted from the UK National Health Service (NHS) perspective, thus only direct costs incurred by the NHS were accounted for. The reference year for costs was 2019. Costs were discounted at 3.5%. Drug costs were sourced from published unit costs such as the British National Formulary (BNF) and the NHS drug tariff and calculated based on posology in the BNF. Biosimilar costs were used where these treatments were available on the NHS. Reasonable assumptions were made regarding confidential discounts, ranging from 25% for innovator biologics to 74% for biosimilars. Costs of biologics were tapered according to NICE guidelines, which recommend withdrawal of biologic after one year of treatment, subject to clinical assessment^13, 18, 19^. Health state costs and costs of flare and surgery were sourced from NICE Health Technology Assessments. Mean values for all cost inputs are provided in Table S1.

### Cost-effectiveness analysis

The outcomes of interest for cost-utility analysis were the differences in outcomes (QALYs) and costs between the two treatment strategies, personalised and standard therapy. This information was combined to derive an incremental cost-effectiveness ratio (ICER). The estimated ICER was compared against the NICE cost-effectiveness threshold of £20,000 to £30,000 per QALY.

### Sensitivity analyses

#### One-way sensitivity analysis

A base-case analysis was undertaken, using a model incorporating the most likely hyperparameters as informed by prior published evidence and independent expert review. Uncertainty in the model results was examined using a one-way sensitive analysis (OWSA) varying key input parameters (Table S2) within their specified range (upper and lower limits). The ICER was recalculated whilst varying one specific parameter and holding all other parameters constant. Results were plotted in the form of a tornado graph, illustrating parameters to which the ICER is most sensitive, quantified as changes in net monetary benefit^1^ (NMB) at the median NICE cost-effectiveness threshold of £25,000 per QALY.

Longer bars indicate the most important parameters, giving the diagram its “tornado” appearance^25^.

#### Scenario analysis

Scenario analyses were conducted to explore the sensitivity of the results to changes in key assumptions underpinning the base-case analysis. The scenarios that were explored were reducing the time horizon, reducing the duration of the treatment effect and reducing the magnitude of treatment effect on clinical outcome (treatment escalations)^6^. A further scenario directly comparing the cost-effectiveness of early top-down versus step-up treatment in the absence of a personalised medicine approach (i.e. without guidance from a prognostic biomarker) was also explored.

#### Probabilistic sensitivity analysis

In addition to OWSA, parameter uncertainty was further explored through probabilistic sensitivity analysis (PSA). PSA involves assigning a distribution to all input parameters, from which samples are repeatedly drawn (5000 iterations in this analysis) and used as model inputs^26^. The purpose of conducting a PSA is to estimate the uncertainty around the ICER value, such that the robustness of the cost-effectiveness estimate can be explored. Output from the PSA is presented in the form of a scatterplot described as the “cost-effectiveness plane”. Depending on the distribution of scatter points within the quadrant(s) of the plane, one can identify the likelihood of the intervention being cost-effective. Points towards the North quadrants indicate greater cost, whereas points towards the East indicate greater likelihood that the intervention is effective^27^. The PSA output was also presented in the form of a cost-effectiveness acceptability curve (CEAC). The CEAC was used to explore the probability of biomarker-guided personalised therapy being cost-effective at a range of ICER thresholds.

### Patient and public involvement

Patient consultation was undertaken during model development through discussion with Crohn’s and Colitis UK.

## RESULTS

### Base-case analysis

A personalised treatment strategy resulted in an ICER of £2,176 per quality-adjusted life year (QALY), with £717 incremental costs and 0.330 incremental QALYs vs. standard of care. The disaggregated costs and outcomes are presented in Table 3. As expected, the costs of induction and maintenance treatment were substantially higher in the personalised therapy group compared to standard care (incremental costs of £9,029 and £112 respectively) but were offset by higher frequency (and therefore costs) of adverse events in the standard care group with respective incremental savings of £1,100 (disease flares), £1,761 (surgery) and £6,812 (hospitalisation costs). Total QALYs were also higher in the personalised therapy group (10.650 vs 10.320), underpinned by higher proportions of patients in better CDAI health states and a reduction in flares and surgery. The results of the fitted survival curves determining the time to switch to the next treatment in the sequence by arm and risk group are depicted in Figures 2A and B. As patients predicted to be ‘low-risk’ in the personalised medicine group receive standard care, they have identical outcomes to patients with a low risk trajectory in the standard care group (orange lines, Figure 2A, B). The key difference in the outcome groups is the lower event rate experienced in high-risk individuals, who receive early aggressive therapy in the personalised group but less effective step-up therapy in the standard care group (orange lines, Figures 2A, B). Assumptions for time to event and the magnitude of benefit brought about by early anti-TNF therapy are based on prior studies as outlined in Table S2 with additional sensitivity modelling as below.

**Table 3:** Base-case scenario results

|  | PredictSURE IBD | No test | Increment |
| --- | --- | --- | --- |
| **Discounted costs** |  |  |  |
| Induction | £4,132 | £4,020 | £112 |
| Maintenance | £33,942 | £24,914 | £9,029 |
| Non-hospitalisation costs | £27,631 | £34,443 | -£6,812 |
| Flares | £1,414 | £2,514 | -£1,100 |
| Surgery and complications | £6,311 | £8,071 | -£1,761 |
| PredictImmune | £1,250 | £0 | £1,250 |
| Total costs | £74,680 | £73,963 | £717 |
| **Clinical outcomes** |  |  |  |
| Number of surgeries per person | 1.183 | 1.513 | -0.330 |
| Life years | 19.554 | 19.549 | 0.005 |
| **Discounted QALYs** |  |  |  |
| QALYs lost to surgery | -0.003 | -0.004 | 0.001 |
| QALYs lost to flares | -0.006 | -0.009 | 0.002 |
| Health state QALYs | 10.659 | 10.332 | 0.326 |
| Total QALYs | 10.650 | 10.320 | 0.330 |

### Sensitivity analysis

#### One-way sensitivity analysis (OWSA)

OWSA was undertaken to determine the impact of independently varying key model assumptions (Figure 3). The model result is illustrated as net monetary benefit (NMB), a summary statistic reflecting the monetary value of an intervention when a willingness to pay threshold (such as QALYs) is known. A positive NMB indicates that an intervention is cost-effective and a value of zero indicating the threshold at which the intervention ceases to be cost-effective for a given QALY threshold^28^. None of the OWSA undertaken reduced the NMB value below, or near to, the cost-effectiveness threshold of £25,000/QALY (NMB £0). The model was most sensitive to the rate of mucosal healing in the TD arm, hospitalisation costs and the odds ratio of steroid-free remission with mucosal healing after 2 years and least sensitive to the observed cost of flares, the proportion of patients receiving treatment for adverse events and the proportion of patients receiving anti-TNF and Immunomodulator treatment due to AEs.

**Figure 3:**
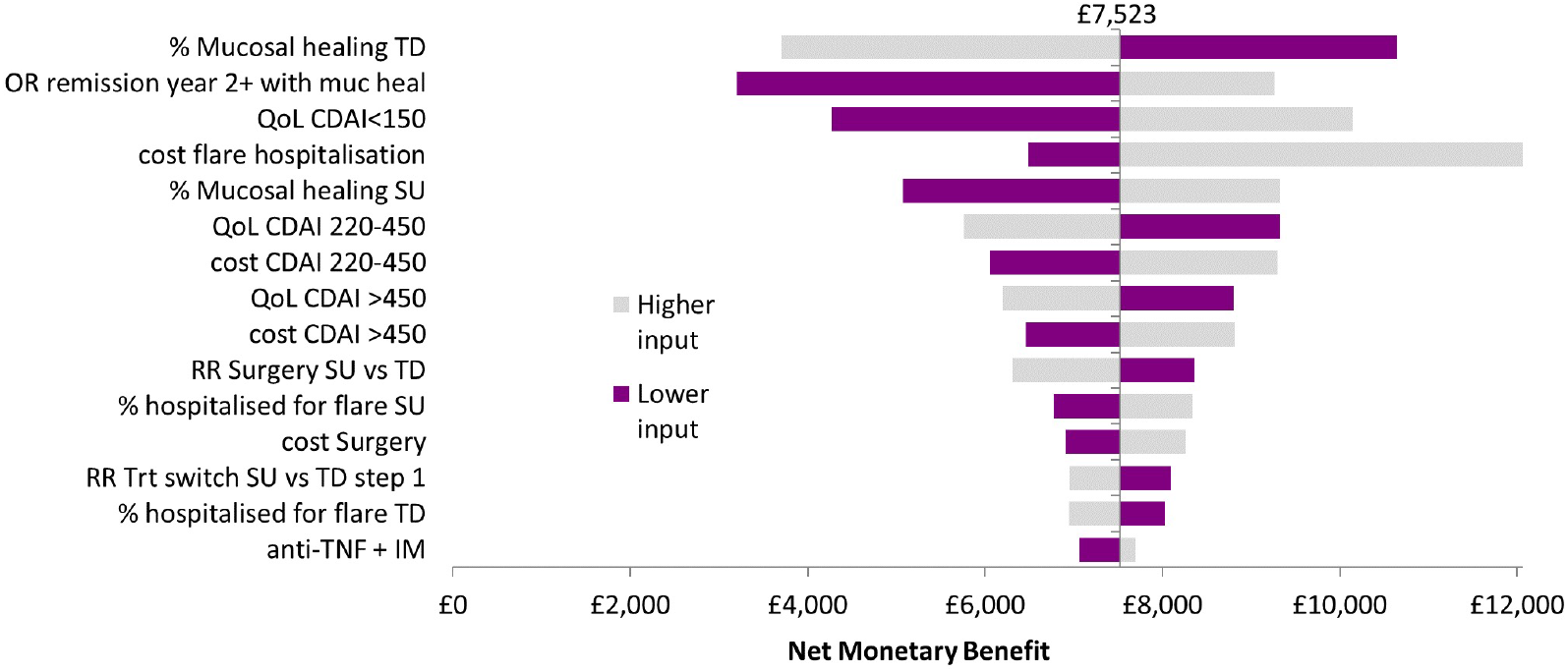
Tornado diagram showing results of varying parameters as part of the one-way sensitivity analysis TD, top-down; SU, step-up; QoL, quality of life; CDAI, Crohn’s disease activity index; RR, relative risk; OR, odds ratio; Trt, treatment. The central line of the tornado indicates the base case NMB of £7,523.

#### Scenario analysis

Reducing the time horizon from 20 years to 15, 10 or 5 years increased the ICER to £7,179,

£26,895 and £99,716 per QALY respectively (Table 4, Figure 5), consistent with increased early costs driven by early adoption of more expensive therapy in the personalised medicine group. There is inevitable uncertainty in estimating the treatment effect size in a personalised medicine strategy: different treatments may be more or less effective when targeted using different prognostic biomarkers. Consequently, we chose to model a wide range of treatment effect size estimates (from 0.3 to 1), ranging from more effective than what has been observed in prior clinical studies of untargeted top down therapy (0.4^6^) to equal effectiveness (Hazard ratio of 1). The cost-effectiveness threshold (NMB £0) was not reached until the effect size was approximately half what has been observed in prior studies (Hazard ratio ∼0.7, Table 5 and Figure 6). A reduction in treatment effect duration from 10 years to 5, 3 or 2 years resulted in an increased ICER of £12,212, £32,625 and £88,307 per QALY respectively (Table 4).

**Figure 4:**
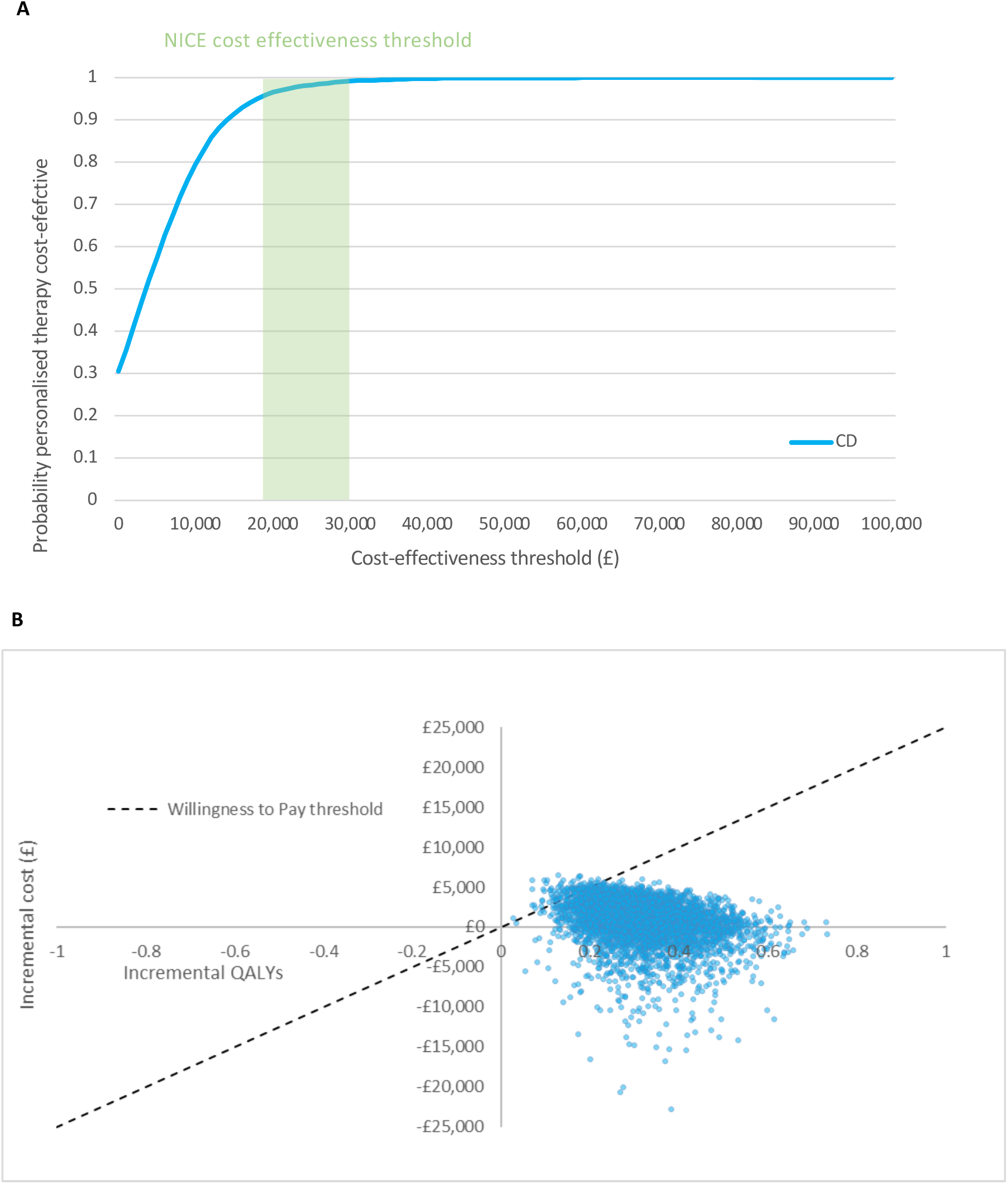
Results of probabilistic sensitivity analysis. **A,** Cost-effectiveness acceptability curve showing the probability of a precision medicine strategy being considered cost-effective at different monetary thresholds; NICE cost-effectiveness upper threshold indicated as green area (£20-30,000/QALY) **B,** Cost-effectiveness plane plotted with scatter points representing 5000 simulations from the probabilistic sensitivity analysis.

**Figure 5:**
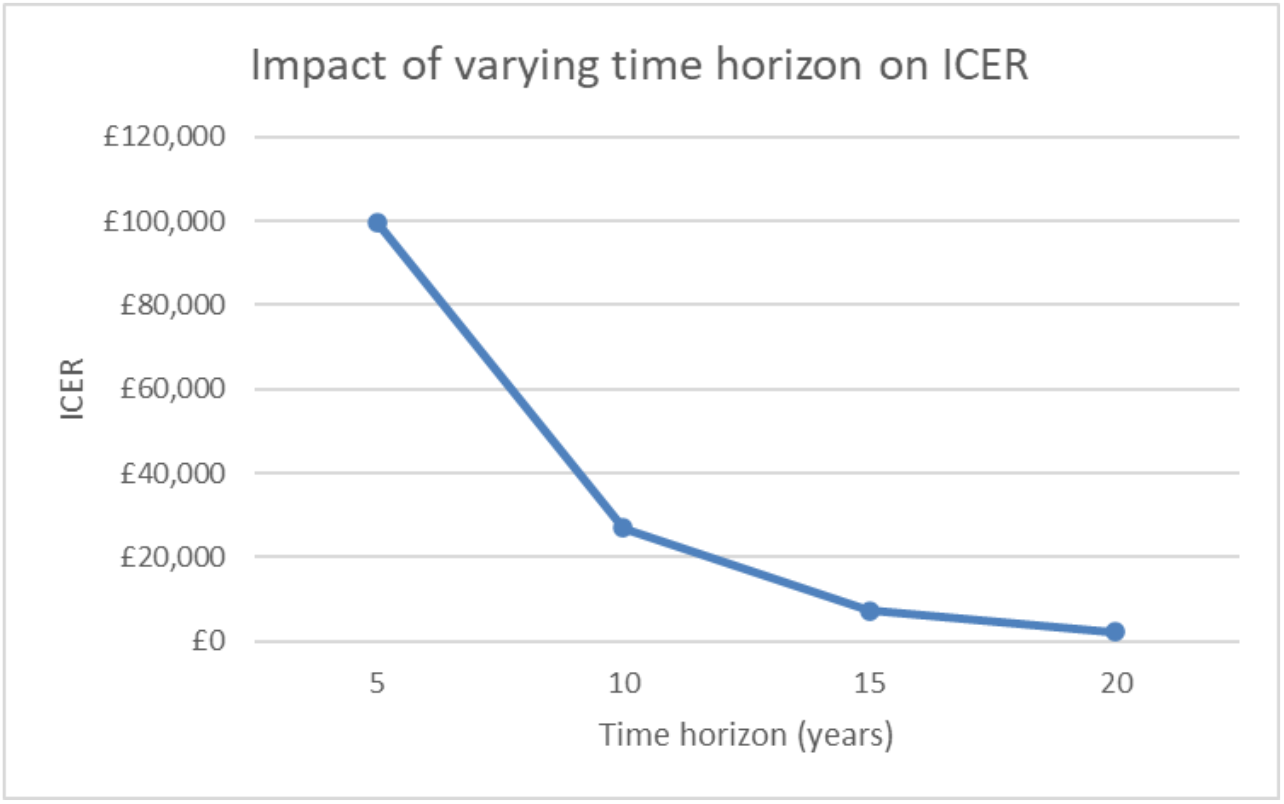
Impact of varying time horizon on cost-effectiveness

**Figure 6:**
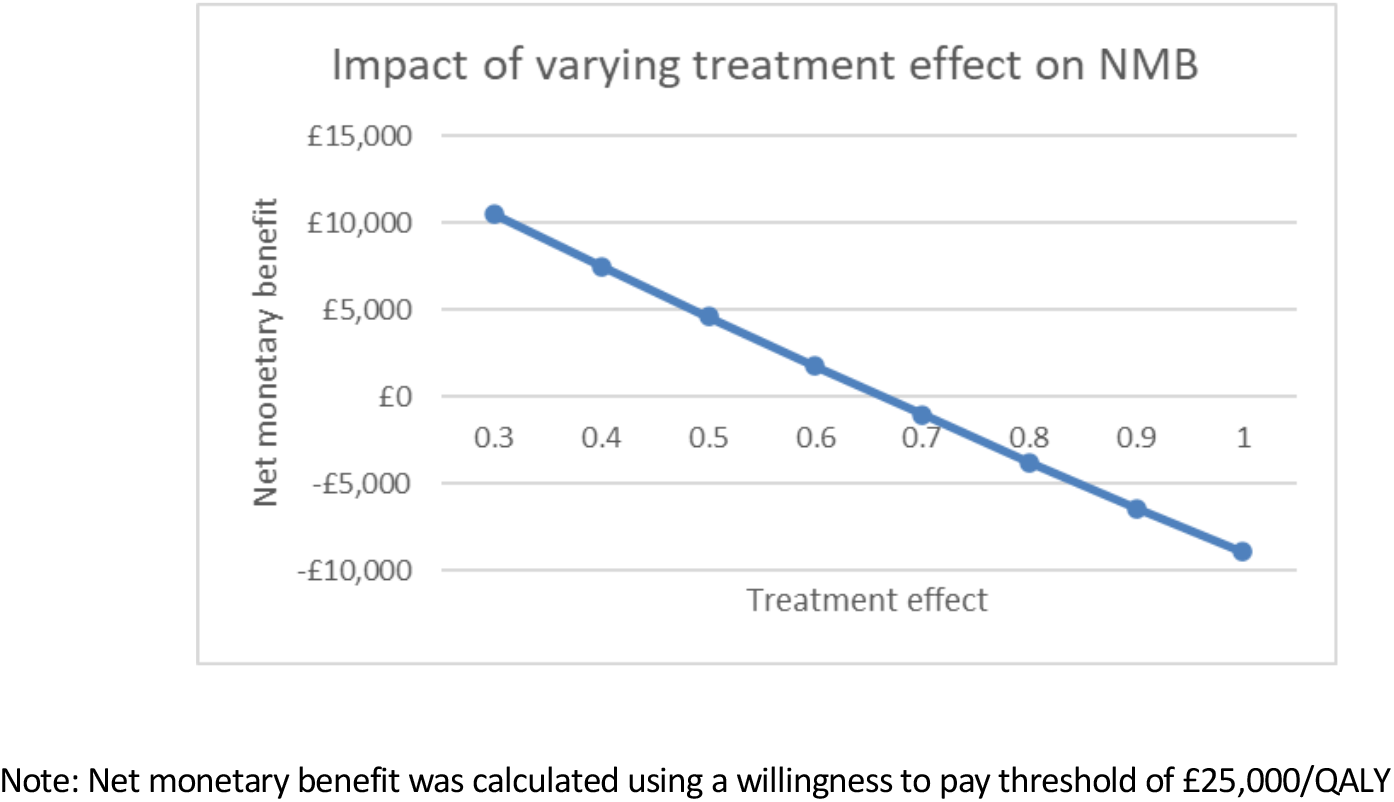
Impact of varying treatment effect size on net monetary benefit

**Table 4:** Scenario analysis results when varying time parameters

| Parameter | Base case value | Value used in scenario analysis | ICER |
| --- | --- | --- | --- |
| Treatment effect on escalations | 0.4 | 0.53 | £9,307 |
| Time horizon | 20years | 15 years | £7,179 |
|  |  | 10 years | £26,895 |
|  |  | 5 years | £99,716 |
| Duration of treatment effect | 10 years | 5 years | £12,212 |
|  |  | 3 years | £32,625 |
|  |  | 2 years | £88,307 |

**Table 5:** Scenario analysis exploring impact on net monetary benefit when varying treatment effect on escalations

| Parameter | Base case value | Value used in scenario analysis | Net monetary benefit |
| --- | --- | --- | --- |
| Treatment effect | 0.4 | 0.3 | £10,475 |
|  |  | 0.4 | £7,523 |
|  |  | 0.5 | £4,633 |
|  |  | 0.6 | £1,780 |
|  |  | 0.7 | -£1,028 |
|  |  | 0.8 | -£3,765 |
|  |  | 0.9 | -£6,399 |
|  |  | 1 | -£8,903 |

A direct comparison of unselected top-down anti-TNF use indicated an ICER of £66,347, with total costs of £74,680 in the ‘all top-down’ arm compared to £93,249 in the standard therapy group (Table 6).

**Table 6:**
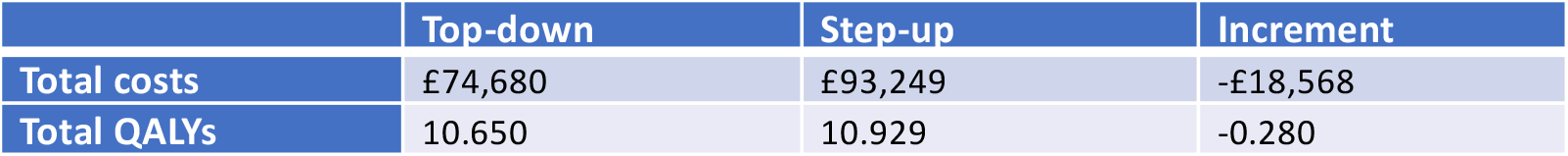
Scenario analysis results for unselected ‘all top down’TD vs standard care

#### Probabilistic sensitivity analysis

A probabilistic sensitivity analysis (PSA) was undertaken to explore the probability of personalised therapy being cost-effective at a range of ICER thresholds. For this approach, across multiple simulations of the input data, an estimate of incremental cost is compared to that of incremental QALYs, allowing an estimate of the probability that an intervention is deemed cost-effective (by falling below a threshold of cost-per-QALY, such as that established by NICE in the UK). This demonstrated that, at the current UK NICE cost-effectiveness threshold of £20-30,000 per incremental QALY, personalised therapy in Crohn’s disease has a ≥95% probability of being cost-effective (cost-effectiveness acceptability curve, Figure 4A). The multiple iterated simulations can be visualised on a cost-effectiveness plane (Figure 4B) which demonstrates that the majority of estimates falling in the upper right (or ‘North East’) quadrant, indicating increased effectiveness with increased costs by adopting a personalised therapy approach (Figure 4B).

## DISCUSSION

The model-based economic evaluation presented here clearly demonstrates that personalised medicine is a cost-effective strategy for CD. The ICER value of £2,176 per QALY is well below the current NICE cost-effectiveness threshold of £20,000 – £30,000 per QALY. As with any model, estimates of likely parameters have been made, using previous high-quality evidence to inform the most likely base-case scenario. A series of on-way sensitivity analyses incorporating broad ranges of all major parameters indicates that the base-case result is unlikely to be altered with modified assumptions.

The incremental QALY gain for the test group was driven by increased time spent in remission and improved QoL as a result of lower rates of flares and subsequent hospitalisations and surgery. A personalised medicine strategy aims to bring more expensive, more aggressive therapy earlier in the treatment paradigm for patients who stand to benefit most from that cost. This inevitably increases early costs compared to a standard step-up regimen. However, the additional costs relating to earlier biologic use in high-risk patients were significantly offset by reductions in the cost of adverse events in these patients. This result is in line with previous economic models although total costs per patient were higher in the present study^8^, driven by increased use of biologic treatments as maintenance (as opposed to purely induction) therapy, consistent with evolving real-world practice. The incremental QALY gain associated with top-down therapy was slightly lower in this study (0.056) compared to that in Marchetti *et al* (2011)^8^ (0.14) as only patients who tested high-risk (41%) received it. While reducing drug costs – due for example to the emergence of biosimilar anti-TNF – could attenuate the increased early expense of a personalised approach, there are inevitably new (and typically comparatively expensive) treatment options that emerge. Consequently, it is unlikely that the model result provided here will substantially change in the near future: reducing drug costs with generic availability are likely to lower the cost of bringing targeted aggressive therapy earlier in the treatment schedule.

Sensitivity analyses undertaken here also indicate the relative importance of key parameter assumptions in determining the cost-effectiveness of a personalised medicine strategy. The ICER was found to reduce significantly with increasing time horizon. Selection of an appropriate time horizon (20 years in the base-case presented here) represents a balance between appropriately capturing the chronic burden of lifelong disease and increasing uncertainty around the persistence of treatment effects (which are typically based on trial evidence with shorter follow up). It is clear from the extended time horizon modelling presented here (Figure 5) that use of very short (5y or less) horizons will inevitably fail to capture later benefit of early, expensive therapy and should be interpreted with caution. As can be expected, use of frontline biologic is more expensive, resulting in higher ICERs when implementing a shorter-term time horizon. When the time horizon is extended, the longer-term benefits of upfront biologic treatment are accrued, resulting in a lower ICER. Notably, NICE prefers the use of longer-term time horizons in chronic diseases, despite the presence of long-term uncertainty from extrapolated outcomes^11^.

The structure of the model applied here is clinically appropriate with the use of treatment sequencing to account for the relapsing-remitting nature of CD. Assumptions were validated with clinician input and key parameters were derived from high quality published evidence with sensitivity analysis to ensure confidence in the model output (Table S2-S4). However, the analysis has some limitations. The cost-effectiveness analysis presented is based on findings from a Markov model. Although demonstrably useful tools in modelling interventions targeted at chronic disease, Markovian models lack ‘memory’ of prior health states and are thus inherently limited for modelling treatment sequencing. We addressed this issue in the present study by assuming constant probabilities for risk of later events and by applying adjustment factors where necessary. QoL outcomes (QALYs) for each health state were also based on published literature and prior NICE technology appraisals^13, 15, 18, 19^. While the model result is sensitive to these QoL inputs (Figure 3) we estimated this uncertainty by varying QoL scores for different health states as part of the OWSA (Figure 3), finding that the overall result remained unchanged.

The current economic evaluation assumes that the prognostic information is available from the point of diagnosis, to guide early ‘top-down’ therapy for patients predicted to follow an aggressive, relapsing disease course. It should be noted that applying a predictive model at later stages of disease, for example by including patients who have already failed on several rounds of ‘step-up’ therapy, is likely to result in smaller treatment effect sizes and less impact on cost-effectiveness. In essence, the later targeted therapy is employed the more the personalised medicine approach resembles a conventional ‘step-up’ regimen.

The extent to which clinical outcomes will be improved by giving targeted anti-TNF therapy to high risk patients at diagnosis remains unknown, although it is being directly tested by the ongoing PROFILE clinical trial^2^. For the current study, assumptions of treatment effect size were informed by the results of the Top-down Step-Up study^6, 23^ and its long-term follow-up extension phase. This study does not directly reflect current clinical approaches to early anti-TNF therapy which have adopted maintenance TNFα inhibition for 1 year in addition to the induction therapy initially trialled. Furthermore, a more extensive range of follow-on biologic therapy options are now available in the case of anti-TNF failure. For this reason we chose to include a marginally higher treatment effect size in our base case compared to that seen in the Top-down Step-Up trial (HR 0.4 vs 0.53), which used infliximab for induction but not maintenance of remission, although it was notable in the sensitivity analysis that a personalised strategy remained cost effective (NMB>£0) until an effect size of almost half the founding estimate was reached (HR 0.7, Figure 6).

## CONCLUSION

This study aimed to evaluate the cost-effectiveness of a personalised medicine strategy for the treatment of newly diagnosed patients with CD, comparing ‘top-down’ use of early anti-TNF therapy with step-up therapy recommended by current NICE guidelines. Our findings suggest that personalised therapy for CD patients is a highly cost-effective strategy with significant implications for the management of CD in the UK.

## Data Availability

All data produced in the present study are available upon reasonable request to the authors

## Author Contributions

James C Lee, Paul A Lyons, Kenneth GC Smith, Miles Parkes and Eoin F McKinney: Conceptualization, Data Curation, Methodology

Vanessa Buchannan, Warda Tahir and Susan Griffin; Conceptualization, Data Curation, Methodology, Formal analysis

Vanessa Buchannan, Warda Tahir, Susan Griffin, Karen Hills & Eoin F McKinney; Writing – original draft and review & editing

James C Lee, Paul A Lyons, Kenneth GC Smith, Miles Parkes; Writing – review & editing

## Data Availability

The data underlying this article are available in this article and the Health Economic model will be shared on reasonable request to the corresponding author.

---

**Table S1:**
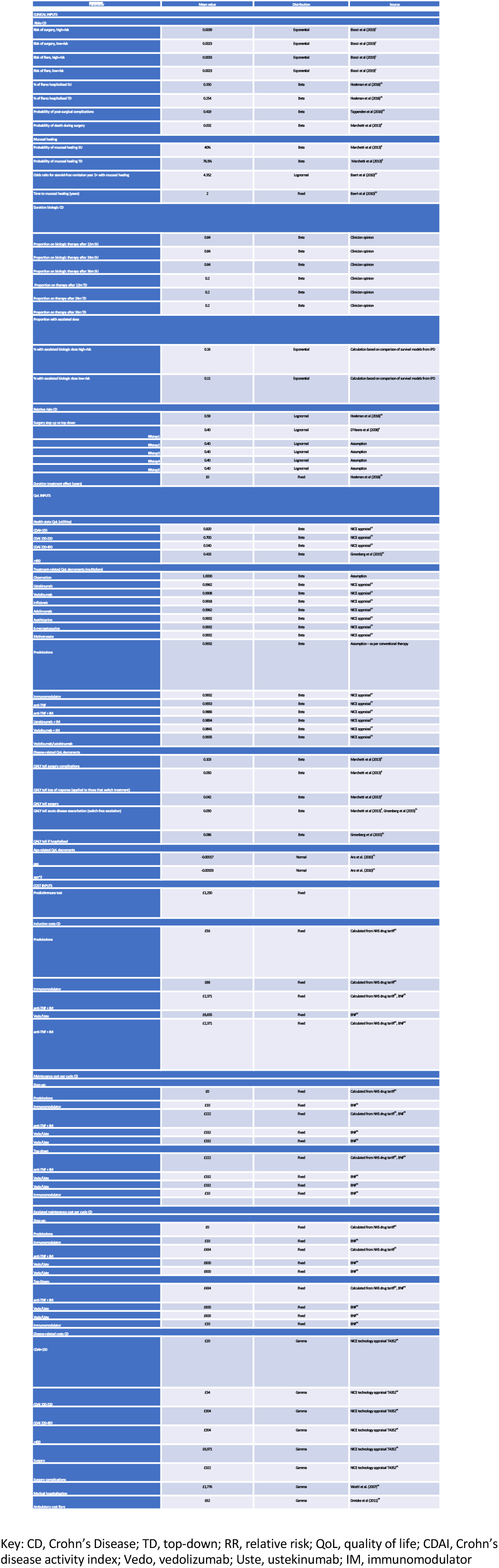
Model input parameters.

**Table S2:** Parameters varied in one-way sensitivity analysis.

| Parameter | Range | Source |
| --- | --- | --- |
| Mucosal healing |  |  |
| Step-up | 0.07 to 0.592 | Marchetti et al (2013)<sup>8</sup> |
| Top-down | 0.55 to 0.923 | Marchetti et al (2013)<sup>8</sup> |
| Hospitalisation costs | 0.64 to 9505 | Woehl et al. (2007)<sup>34</sup> |
| Surgery costs | 4511 to 9128 | NICE technology appraisal TA352<sup>19</sup> |
| Cost of treating flare | 0.1639 to 289 | Dretzke et al (2011)<sup>35</sup> |
| OR steroid-free remission year 2+ with mucosal healing | 1.1 to 17.2 | Baert et al (2010)<sup>20</sup> |
| Relative risk of surgery step-up vs top-down | 0.41 to 0.65 | Hoekman et al (2018)<sup>23</sup> |
| Relative risk of treatment switch (step-up vs top-down) from induction to maintenance treatment | 0.37 to 0.43 | D'Haens et al (2008)<sup>6</sup> |
| Hospitalised for Flare StepUp | 0.24 to 0.47 | Hoekman et al (2018)<sup>23</sup> |
| Hospitalised for Flare Top-Down | 0.15 to 0.37 | Hoekman et al (2018)<sup>23</sup> |
| Cost of CDAI <150 | 13 to 28 | NICE technology appraisal TA352<sup>19</sup> |
| Cost of CDAI 150-220 | 22 to 48 | NICE technology appraisal TA352<sup>19</sup> |
| Cost of CDAI 220-450 | 197 to 434 | NICE technology appraisal TA352<sup>19</sup> |
| Cost of CDAI >450 | 197 to 434 | NICE technology appraisal TA352<sup>19</sup> |
| Utility of CDAI <150 | 0.71 to 0.91 | NICE technology appraisal TA352<sup>19</sup> |
| Utility of CDAI 150-220 | 0.66 to 0.74 | NICE technology appraisal TA352<sup>19</sup> |
| Utility of CDAI 220-450 | 0.40 to 0.68 | NICE technology appraisal TA352<sup>19</sup> |
| Utility of CDAI >450 | 0.30 to 0.57 | Marchetti et al (2013)<sup>8</sup> |
| Utility multiplier due to Adverse Event: |  |  |
| Biologic (Vedolizumab/Ustekinumab) | 0.98 to 1 | NICE technology appraisal TA352<sup>19</sup> |
| anti-TNF + IM | 0.96 to 1 | NICE technology appraisal TA352<sup>19</sup> |
| Immunomodulator | 0.97 to 1 | NICE technology appraisal TA352<sup>19</sup> |

**Table S3:** Clinical cohort characteristics by risk subgroup (after Biasci et al, Gut 2019)

|  |  |  |  |
| --- | --- | --- | --- |
|  | high risk | low risk | P value |
| Age (years) | 30.3 (25.3-36.1) | 30.3 (23.2-38.7) | 0.98 |
| Sex (% male) | 42.4 | 39.4 | 1 |
| smoking (% yes) | 28.6 | 33.3 | 0.79 |
| newly diagnosed (% yes) | 81.8 | 72.7 | 0.56 |
| disease duration (years) | 0 | 0 | 0.78 |
| Haemoglobin (g/L) | 12.5 (11.7-13.3) | 13.1 (11.8-13.6) | 0.63 |
| CRP (mg/L) | 26 (16-39) | 25 (10-59) | 0.6 |
| Albumin (g/L) | 35 (32-37) | 37 (34-39) | 0.14 |
| Disease distribution: |  |  |  |
| ileal | 27.3 | 39.4 | 0.43 |
| ileocolonic | 33.3 | 27.3 | 0.79 |
| colonic | 39.4 | 33.3 | 0.8 |
| upper GI | 6.1 | 9.1 | 1 |
| perianal | 6 | 3 | 0.48 |
| prospective follow-up, years | 4.9 (3.6-7.4) | 5.3 (4.3-8.3) | 0.24 |

**Table S4:** Model validity evaluation criteria. Following ISPOR recommendations as per Principles of Good Practice for Decision Analytic Modeling in Health, Value Health. 2003;6(1):9-17. doi:10.1046/j.1524-4733.2003.00234

| Model Criterion | Criterion met? |  |
| --- | --- | --- |
|  | Yes | No |
| **A. Model structure** |  |  |
| inputs/outputs relevant to decision-making perspective | x |  |
| evidence supporting causal linkages between variables | x |  |
| limitations accepted (sensitivity analyses performed) | x |  |
| Health states defined to correspond to disease process | x |  |
| State transition probabilities depend on prior states | x |  |
| States not excluded due to lack of data | x |  |
| Consideration given to health state subdivision | x |  |
| appropriately short cycle length | x |  |
| model structure simplicity | x |  |
| model options not constrained by availability of evidence | x |  |
| data availability may affect model choices | x |  |
| disaggregation of founding population | N/A |  |
| time horizon long enough to reflect long term consequences | x |  |
| **B. Data input** |  |  |
| **i. Data identification** |  |  |
| literature reviews conducted on key inputs | x |  |
| ranges for parameters used in sensitivity analysis | x |  |
| formal probabilistic sensitivity analysis | x |  |
| justification of any excluded data sources | N/A |  |
| uncertainty around estimates disclosed | x |  |
| sensitivity analysis for expert opinion input | x |  |
| reasonable opportunities to include new data included | x |  |
| **ii. Data modeling** |  |  |
| assumptions disclosed and supported by evidence | x |  |
| sensitivity analyses with alternative modeling if appropriate | N/A |  |
| modeling following accepted principles | x |  |
| **iii. Data incorporation** |  |  |
| measurement units, time intervals and populations consistent | x |  |
| probabilistic or deterministic simulation | x |  |
| probabilistic or deterministic sensitivity analyses for key parameters | x |  |
| half-cycle correction used where appropriate | N/A |  |
| **C. Model validation** |  |  |
| **i. internal validation** |  |  |
| model debugging e.g. using extreme values and sense checking | x |  |
| model calibration against external data | x |  |
| availability of model on reasonable request | x |  |
| **ii. Between model validation** |  |  |
| model developed independently | x |  |
| discrepancies with published models explained | x |  |
| cooperation with other modelers for comparison/explanation where appropriate | x |  |
| **iii. External and predictive validation** |  |  |
| results reported as conditional on the input assumptions | x |  |
| consideration given to obtaining additional prospective data for validation | x |  |
| models open to update or replacement when new data becomes available | x |  |

## Footnotes

1 calculated as incremental QALYs * willingness to pay threshold – incremental costs

